# Sodium and Potassium Consumption in Jamaica: National Estimates and Associated Factors from the Jamaica Health and Lifestyle Survey 2016-2017

**DOI:** 10.1101/2023.02.18.23286134

**Authors:** Trevor S Ferguson, Novie Om Younger-Coleman, Karen Webster-Kerr, Marshall K. Tulloch-Reid, Nadia R Bennett, Tamu Davidson, Andriene S Grant, Kelly-Ann M. Gordon-Johnson, Ishtar Govia, Suzanne Soares-Wynter, Joette A Mckenzie, Evelyn Walker, Colette A Cunningham-Myrie, Simon G Anderson, Alphanso L Blake, James Ho, Robyn Stephenson, Sharmaine E Edwards, Shelly R Mcfarlane, Simone Spence, Rainford J Wilks

## Abstract

**Objective:** To estimate dietary sodium and potassium consumption among Jamaicans and evaluate associations with sociodemographic and clinical characteristics.

**Methods:** We conducted a cross-sectional analysis of data from the Jamaica Health and Lifestyle Survey 2016-2017. Participants were non-institutionalized Jamaicans, ≥15 years. Trained staff collected sociodemographic and health data via interviewer administered questionnaires and collected spot urine samples. The Pan American Health Organization Formulae were used to estimate 24-hour urine sodium and potassium excretion. High sodium was defined as ≥2000 mg/day and low potassium as <3510 mg/day (World Health Organization criteria). Associations of these outcomes with sociodemographic and clinical characteristics were explored in sex specific multivariable ANOVA models.

**Results:** Analyses included 1009 participants (368 males, 641 females; mean age 48.5 years). Mean sodium excretion was 3582 mg/day (males 3943 mg/day, females 3245 mg/day, p<0.001). Mean potassium excretion was 2052 mg/day (males 2210 mg/day, females 1904 mg/day, p=0.001). The prevalence of high sodium consumption was 66.6% (males 72.8%, female 60.7%, p<0.001) and low potassium intake was 88.8% (85.1% males, 92.3% females, p<0.001). Among males, sodium consumption was inversely associated with older age and prehypertension, but directly associated with current smoking and obesity. Among females, sodium consumption was inversely associated with hypertension, impaired fasting glucose, low GFR and high physical activity, but was directly associated with obesity.

**Conclusion:** Most Jamaican adults have diets high in sodium and low in potassium. Sodium consumption was directly associated with obesity in both men and women. Population based strategies are therefore required to address these cardiovascular risk factors.

## INTRODUCTION

Excess sodium consumption is a major risk factor for hypertension and cardiovascular diseases (CVD) and has been estimated to cause between three to five million deaths globally each year (1-3). While the human body needs small amounts of sodium to maintain homeostasis, excess sodium has been associated with several adverse health outcomes, including high blood pressure, heart disease, stroke, chronic kidney disease, stomach cancer and obesity (1). The World Health Organization (WHO) recommends that sodium intake be limited to < 2.0 g per day, but the global average for sodium intake far exceeds that, at approximately 3.95 g per day (4, 5). Given this high sodium intake, the WHO has published recommendations for member states to institute measures to reduce dietary sodium intake both at the individual and population level (5). The population approach as a strategy to achieve sodium reduction is also recommended by the Pan American Health Organization (PAHO) and the Caribbean Public Health Agency (CARPHA) (6, 7).

There is strong evidence to support the potential impact of dietary salt reduction in the general population as a strategy for the reduction of hypertension and CVD (8-14). An example of the successful implementation of population wide sodium reduction was seen in the United Kingdom (UK), where a programme of voluntary salt reduction was developed and implemented in collaboration with the food industry in 2003 to 2004 (11, 15). This resulted in a 15% reduction in mean population salt intake, from 9.5 g daily in 2003 to 8.1 g daily in 2011 (10, 11). Research later demonstrated a 2.9 mm Hg reduction in the population mean systolic blood pressure after accounting for potential confounding factors (10, 11). They also found that stroke and ischaemic heart disease (IHD) mortality were reduced by 36% over the same period (10). Successful population wide salt reduction programmes have been demonstrated in other countries, including Finland, Japan and Portugal (10).

Potassium intake is associated with several health outcomes, including hypertension and cardiovascular diseases (16, 17). Most studies have found that higher potassium intake was associated with lower blood pressure (16, 17). It has also been found that the ratio of sodium to potassium intake is also strongly associated with hypertension and CVD (18). WHO recommends an increase in potassium intake from food to reduce blood pressure and risk of cardiovascular disease, stroke and coronary heart disease in adults (19). WHO suggests that adults have a potassium intake of at least 90 mmol/day (3510 mg/day) (19).

Cardiovascular disease and hypertension are major health problems in Jamaica, with hypertension being consistently among the top five leading causes of death (20, 21). Data from the most recent Jamaica Health and Lifestyle Survey conducted in 2016-2017 (JHLS-III) showed that two-thirds of the population have an abnormally high blood pressure; approximately one third (33.8%) had hypertension (blood pressure more than 140/90 mm Hg) and 34.0% had prehypertension (systolic blood pressure 120-139 mm Hg or diastolic blood pressure of 80-89 mm Hg) using the JNC-7 classification (22).

There are very little data on salt consumption patterns among Jamaicans, but it is generally believed that salt consumption is high. The Spanish Cohort Study reported estimated sodium excretion of 3.3 g/24 hour in the 1990s (23), we do not have any recent or national estimates for Jamaica. Development and implementation of programmes, including salt reduction strategies, are critical actions if Jamaica is to mitigate the impact of elevated blood pressure on population health; however adequate baseline data are needed to inform policies and assess their impact. In this study, we estimate dietary sodium and potassium consumption among Jamaicans using spot urinary analyses and conduct exploratory analyses to evaluate associations between sodium and potassium consumption and sociodemographic and clinical characteristics.

## METHODS

### Study design and Data Sources

We conducted a cross-sectional analysis of data collected as part of the Jamaica Health and Lifestyle Survey 2016-2017 (JHLS-III), a national health examination survey conducted between September 2016 and February 2017. Details of the procedures used in JHLS-III and the protocol for this study have been published previously (24, 25). In brief, trained personnel were engaged to collect data on a wide cross-section of health-related indices using interviewer-administered questionnaires, physical measurements, point-of-care and laboratory measurements and geographic information system (GIS) mapping. A multiple-stage sampling design was used to recruit participants 15 years and older. A total of 171 enumeration districts (EDs) were selected based on probability proportionate to the size of the ED. Within each ED 20 households (dwellings) were systematically selected beginning at a random starting point. Within each household one individual meeting the inclusion criteria was selected using the Kish method (26). Both JHLS-III and this study were reviewed and approved by the Ethics Committees of the University of the West Indies (UWI) - UWI – ECP 25, 15/16, - and the Ministry of Health and Wellness (MOHW) - CREC-MN.153 2020/2021; MOHW – 2015/51, 2021/05. All participants provided written informed consent.

### Questionnaire Data

An interviewer administered questionnaire was used to collect sociodemographic data, medical history, including diagnosis and medication use for conditions such as diabetes and hypertension, and information on health behaviours including dietary practices, physical activity, and tobacco smoking.

Educational level was categorized based on participants’ responses to a question on the highest level of education reached. Categories were: ‘less than high school’ for persons reporting no education, elementary/primary school education, or junior secondary school education (up to grade 8), ‘high school’ for participants reporting at least some secondary level education (grades 9-13) and ‘more than high school’ for those reporting at least some post-secondary school education, whether through college, university, or vocational training institutions.

Household socioeconomic status was assessed based on participants’ responses to whether they had a list of 22 household assets. The full list of items is show in Table S1 of the supplementary file. We created tertiles of the number of household assets for use in the analyses. Tertile 1 included persons with nine or fewer household assets, tertile 2 included those with 10-12 items and tertile 3 persons with 13-22 items.

Physical activity was estimated from the International Physical Activity Questionnaire [IPAQ] short form, and physical activity levels were classified as low moderate or high using cut recommended by the IPAQ developers (27).

Tobacco smoking was assessed using the question: “ *Do you currently smoke any form of tobacco (cigarettes, cigars, beady etc*.*)?”* Participants who answered yes to this question were categorized as current smokers.

### Blood pressure and anthropometric measurements

Blood pressure was measured using an oscillometric device (Omron 5 series blood pressure monitor, Omron Healthcare, Lake Forest, IL). Three measurements were obtained from the right arm of seated participants after ensuring at least five minutes of rest. These procedures were developed and standardized for the International Collaborative Study of Hypertension in Blacks (28). To ensure reliability of measurements, field staff were trained and certified prior to starting fieldwork and at approximately three-month intervals thereafter. The means of the second and third systolic blood pressure (SBP) and diastolic blood pressure (DBP) measurements were used in the analyses. Blood pressure was categorized using criteria from the Seventh Report of the Joint National Committee on Prevention, Detection, Evaluation, and Treatment of High Blood Pressure (29). Normal blood pressure was defined as SBP <120 mm Hg and DBP <80 mm Hg; persons with SBP 120-139 mm Hg or DBP 80-89 mm Hg were classified as prehypertension; hypertension was defined as SBP ≥ 140 mm Hg or DBP ≥ 90 mm Hg or participant report of previously being on medication for hypertension.

Weight was measured using a portable digital scale (Tanita HD-351 Digital Weight Scale, Tanita Corporation, Tokyo, Japan) and recorded to the nearest 0.1 kg. Height was measured using a portable stadiometer (Seca 213 Mobile stadiometer, Seca GmbH & Co., Hamburg, Germany) and recorded to the nearest 0.1 cm. Body mass index (BMI) was calculated as the weight in kilograms divided by the square of height in metres. BMI was categorized using World Health Organization (WHO) criteria (30). Persons with BMI <18.5 kg/m^2^ were classified as underweight; normal was defined as BMI 18.5 – 24.99 kg/m^2^; overweight defined as BMI 25.0 – 29.99 kg/m^2^ and obese as BMI ≥ 30.0 kg/m^2^.

### Point of Care and Laboratory Measurements

Fasting glucose and total cholesterol were measured from a capillary blood sample using a point of care device (SD LipidoCare, Suwon, South Korea). Diabetes cut points were based on the American Diabetes Association criteria (31), with diabetes defined as fasting glucose ≥ 7.0 mmol/l or participants’ report of previously being on medications for diabetes; impaired fasting glucose was defined as fasting glucose of 5.6 to 6.9 mmol/l for persons without a previous history of being on medications for diabetes mellitus and normal glucose defined as fasting glucose <5.6 mmol and no previous history of being on medications for diabetes mellitus. Cholesterol cut points were based on Adult Treatment Panel III (32), with high cholesterol defined as fasting total cholesterol ≥ 5.2 mmol/l or participants report of being on medications for high cholesterol.

Serum creatinine was measured using the Cobas c111 Analyzer (Roche Diagnostics, Basel, Switzerland) and reported in µmol/l. Estimated glomerular filtration rate was obtained from serum creatinine using the formula from the Chronic Kidney Disease Epidemiology (CKD-EPI) Collaboration (33) and reduced GFR defined as eGFR <60 ml/min/1.73 m^2^ (34).

### Estimated 24-hour urine sodium and potassium excretion

An early morning spot urine sample was collected from study participants and urine sodium and potassium concentrations were measured using the Roche 9180 Electrolyte Analyzer (Mannheim, Germany). Urine creatinine was measured using the Cobas c111 Analyzer (Roche Diagnostics, Basel, Switzerland). Spot urine sodium and potassium concentrations were used to estimate 24-hour urine sodium (Na) and potassium (K) levels using formulae recommended by PAHO shown in Equation 1 below (35). Estimated 24-hour urine creatinine was calculated using the Chronic Kidney Disease Epidemiology Collaboration (CKD-EPI) formula (Equation 2) shown below (36, 37):

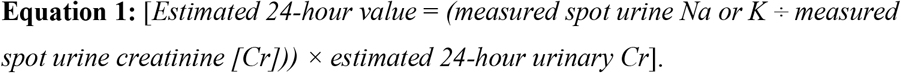

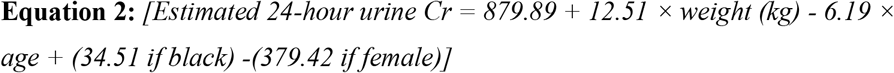

We then used the 24-hour urine sodium and potassium levels to categorize participants’ sodium and potassium consumption categories, and to estimate sodium to potassium ratio. High sodium consumption was defined as ≥ 2000 mg/day while low potassium consumption was defined as < 3510 mg/day according to WHO criteria (5, 19, 38). Normal sodium consumption was defined as <2000 mg/day, while adequate potassium consumption was defined as ≥3510 mg/day. High sodium to potassium ratio was defined as ≥ 1.0 as recommended by the INTERSALT investigators (18).

### Sample size and power

Details of sample size and power calculations were reported in the study protocol (25). We had urine samples from 1,091 participants which translated to an effective sample size of 563 participants after applying a design effect of 1.94 to account for the survey design. Hypothesized mean sodium excretion of 2,565 mg per day was obtained from the Health of the Nations Study in Barbados (39). Using the ‘*power onemean’* command in <u>Stata</u> (RRID:SCR_012763) and 10% margin of error (*i*.*e*., 2,656 vs. 2,390), the effective sample size of 563 gave a power of 97.5% for this study.

### Statistical analyses

Data were analyzed using Stata version 16.1 (StataCorp, College Station, Texas). We estimated means and proportions for 24-hour urine sodium, 24-hour urine potassium and related variables and assessed associations with covariates including age, sex, socioeconomic status, health behaviour and clinical characteristics. Differences in proportions were estimated using Pearson’s chi-squared test or Fishers exact test, while differences in means were estimated using Students t-test or analysis of variance, as appropriate. Analyses for population mean sodium and potassium consumption, estimates for prevalence of high sodium and low potassium, bivariate models and multivariable models were done using survey weights to account for the multi-stage survey design and the age and sex distribution of the population. Descriptive statistics showing sample means and proportions were unweighted. Prevalence of high sodium consumption and low potassium consumption were estimated within and across sex and age categories. The distributions for both 24-hour urine sodium and 24-hour urine potassium were right skewed and therefore were log-transformed for use in bivariate and multivariable analysis of variance (ANOVA) models. There was evidence for sex interaction in the relationship between sodium and potassium consumption and some explanatory variables, therefore we present sex-specific bivariate and multivariable models. These sex-specific multivariable ANOVA models were treated as our final (primary) analytic results.

Covariates included in developing the multivariable analyses were age category (in 20-year bands), sex, socioeconomic status (education level and number of household possessions), adding salt at the table, cigarette smoking and physical activity, blood pressure category, diabetes status, BMI categories, high cholesterol and low eGFR. Those selected for inclusion in the final models had at least one category with a p-value <0.2 for association with the outcome in the bivariate models. We also estimated prevalence ratios for high sodium and low potassium consumption using Poisson regression models. We chose to report prevalence ratios rather than odds ratios, given that in situations where the prevalence of the outcome is high the odds ratio may overestimate the relative risk estimate (40, 41).

Analyses were restricted to participants who had non-missing data 24-hour urine sodium, age, sex, and survey weights resulting in an analytic sample of 1009 participants. Of these 1009 included participants, 510 (50.5%) had no missing values for any of the variables selected for use in the analyses; 499 participants (49.5%) had one or more missing values. The missing data were due primarily to smoking with 34% of participants having missing data; eight percent of participants had missing data for fasting cholesterol and six percent had missing data for fasting glucose. Proportion of missing data was less than 5% for all other variables with missing data. There were no statistically significant differences between participants with missing data and those without missing data when we compared the means and proportions for variables used in the analyses. Table S2 in the supplementary files shows the number and proportion of missing values for the variables included in the analyses. Given the proportion of participants with missing data, complete case analysis (list-wise deletion of missing data) would result in loss of power, therefore we chose to use multiple imputation to deal with missing data in this paper (42, 43). We generated 50 imputed datasets, such that the number of imputations would be at least equal to the proportion of incomplete cases, representing an adaptation of a rule of thumb suggested by White and colleagues (43). Imputation models included all the variables for inclusion in multivariable models. Bivariate and multivariable models were created using Stata’s *mi estimate* command using the fifty multiply imputed data sets; estimates were combined by the software using Rubin’s rules (44, 45). We also performed complete case analyses (i.e., model with no imputed values) to assess whether inferences were similar to those obtained from the multiple imputation analyses.

## RESULTS

The final analysed sample included 1009 participants (368 males, 641 females). Summary statistics for participant characteristics are shown in Table 1, Mean age of participants was 48.5 ± 18.2 years, with no sex difference. Males were taller than females, but females had higher mean weight and higher mean BMI (30.2 kg/m^2^ vs 24.7 kg/m^2^, p<0.001). Males also had higher mean SBP, but women had higher fasting glucose cholesterol. Mean DBP and eGFR were similar for men and women. Mean spot urine sodium concentration was 2973 mg/l and was higher in men compared to women (3151 mg/l vs 2870 mg/l, (p=0.001)). Mean spot urine potassium concentration was 2319 mg/l and was not significantly different between sexes. Survey weighted estimates are shown in Table S3 in the supplementary file. Overall mean age was lower than the unweighted estimates at 40.1 years (95% CI 40.0, 40.2). There was also a slightly higher mean spot urine sodium concentration of 3150 mg/l and slightly lower spot urine potassium concentration of 2119 mg/l.

**Table 1:** Summary statistics (means and standard deviations) for participant characteristics within and across sex categories.

| Characteristic | Male<br>n = 368<br>Mean ± SD | Female<br>n = 641<br>Mean ± SD | Total<br>N = 1009<br>Mean ± SD | p-value for<br>Male:<br>Female<br>differences |
| --- | --- | --- | --- | --- |
| Age (years) | 49.3 ± 19.1 | 48.1 ± 17.7 | 48.5 ± 18.2 | 0.325 |
| Weight (kg) | 74.1 ± 21.8 | 79.1 ± 23.9 | 77.3 ± 23.3 | <0.001 |
| Height (cm) | 171.7 ± 12.9 | 161.5 ± 13.0 | 165.3 ± 13.9 | <0.001 |
| Body mass index (kg/m<sup>2</sup>) | 24.71 ± 6.8 | 30.2 ± 8.9 | 28.2 ± 8.6 | <0.001 |
| Systolic Blood Pressure (mmHg) | 133.0 ± 21.1 | 129.6 ± 22.8 | 130.9 ± 22.3 | 0.018 |
| Diastolic Blood Pressure (mmHg) | 83.0 ± 13.2 | 83.8 ± 12.1 | 83.5 ± 12.5 | 0.351 |
| Fasting glucose (mmol/l) | 5.66 ± 1.32 | 6.15 ± 2.74 | 5.98 ± 2.35 | <0.001 |
| Total Cholesterol (mmol/l) | 4.42 ± 1.02 | 4.62 ± 1.04 | 4.55 ± 1.03 | 0.005 |
| Estimated GFR (ml/min/1.73m<sup>2</sup>) | 95.2 ± 26.4 | 97.0 ± 27.4 | 96.3 ± 27.1 | 0.332 |
| Spot urine sodium (mg/l) | 3150.5 ± 1314.2 | 2870.4 ± 1308.7 | 2972.5 ± 1317.0 | 0.001 |
| Spot urine potassium (mg/l) | 2036.5 ± 1151.6 | 2197.5 ± 1888.3 | 2139.0 ± 1660.0 | 0.094 |
| 24-hour urine creatinine (CKD-EPI, mg) | 1534.8 ± 304.2 | 1225.8 ± 321.9 | 1338.5 ± 348.8 | <0.001 |

Estimates derived from unweighted analyses; P-values for differences in means obtained from t-tests. CKD-EPI = Chronic Kidney Disease Epidemiology Collaboration.

Proportions for participants’ biomedical and socioeconomic characteristics are shown in Table 2. Only 31% of participants had normal blood pressure; 38% were classified as prehypertensive and 32% of participants were hypertensive and. There was evidence of a significant sex difference in the distribution of blood pressure categories (p<0.001), with a higher proportion of men having prehypertension and higher proportion of women having normal blood pressure. Sex differences were also recorded in the BMI and physical activity categories (p<0.001 for both). A higher proportion of women was found to be obese (41.7% vs 18.5%), while men were more likely to be underweight (7.4% vs. 1.7%). Physical activity levels also varied significantly between sexes (p<0.001), with more women reporting low physical activity levels (45% vs 24%) and more men reporting high physical activity levels (52% vs 25%). Additionally, while 17% of the sample reported current smoking r, significantly more men than women reported smoking (27.7% vs 7.6% (p<0.001)). Most participants (58%) had attained secondary education, with a slightly higher proportion of women attaining tertiary education (18% vs 13%).

**Table 2:** Proportion (%) of Participant Characteristics within and across sex categories.

| Characteristic | Male<br>n = 368<br>% (95% CI) | Female<br>n = 641<br>% (95% CI) | Total<br>N = 1009<br>% (95% CI) |
| --- | --- | --- | --- |
| Blood Pressure Category *** |  |  |  |
| Normal BP | 24.4 (21.3, 28.0) | 36.6 (32.7, 40.6) | 30.7 (28.2, 33.3) |
| Prehypertension | 47.0 (42.7, 51.4) | 28.7 (25.5, 32.1) | 37.6 (34.8, 40.4) |
| Hypertension | 28.4 (24.8, 32.4) | 34.7 (31.5, 38.0) | 31.7 (29.5, 34.0) |
| Blood Glucose Level |  |  |  |
| Normal | 58.7 (54.2, 63.2) | 56.0 (52.1, 59.8) | 57.3 (54.4, 60.2) |
| Impaired Fasting Glucose | 31.0 (26.4, 36.0) | 29.3 (25.4, 33.4) | 30.1 (27.3, 33.0) |
| (ADA) Diabetes | 10.3 (7.5, 14.0) | 14.8 (12.4, 17.5) | 12.6 (10.7, 14.8) |
| High Cholesterol | 18.8 (16.0, 21.8) | 21.2 (18.8, 23.8) | 20.0 (18.0, 22.3) |
| Body Mass Index categories *** |  |  |  |
| Normal (18.5 – 24.99 kg/m<sup>2</sup>) | 46.0 (42.3, 49.7) | 31.9 (28.5, 35.5) | 38.7 (36.1, 41.3) |
| Underweight (< 18.5 kg/m<sup>2</sup>) | 7.4 (5.2, 10.4) | 1.7 (0.9, 3.1) | 4.4 (3.4, 5.7) |
| Overweight (25 – 29.99 kg/m<sup>2</sup>) | 28.2 (25.0, 31.5) | 24.7 (21.7, 28.0) | 26.4 (24.4, 28.4) |
| Obese (≥ 30 kg/m<sup>2</sup>) | 18.5 (14.9, 22.7) | 41.7 (38.0, 45.6) | 30.5 (28.2, 33.0) |
| Physical Activity Level *** |  |  |  |
| Low | 23.7 (20.4, 27.4) | 45.1 (41.6, 48.6) | 34.7 (32.1, 37.5) |
| Moderate | 24.8 (21.0, 29.0) | 30.3 (26.7, 34.2) | 27.6 (24.8, 30.7) |
| High | 51.5 (47.1, 55.9) | 24.6 (21.5, 28.0) | 37.6 (34.6, 40.7) |
| Education * |  |  |  |
| Less than High School | 28.7 (25.8, 31.8) | 23.5 (20.6, 26.6) | 26.0 (24.2, 27.9) |
| High School | 58.8 (54.0, 63.4) | 58.2 (53.8, 62.6) | 58.5 (55.5, 61.4) |
| More than High School | 12.5 (9.8, 15.8) | 18.3 (14.5, 22.9) | 15.5 (13.0, 18.4) |
| Household Possessions |  |  |  |
| ≤8 items | 35.3 (31.8, 39.0) | 32.5 (29.1, 36.1) | 33.9 (31.5, 36.3) |
| 9-11 items | 32.6 (28.7, 36.8) | 34.0 (29.7, 38.5) | 33.3 (30.6, 36.1) |
| 12-22 items | 32.0 (28.5, 35.9) | 33.5 (29.4, 37.9) | 32.8 (29.7, 36.1) |
| Low GFR | 3.8 (3.0, 4.9) | 4.8 (3.7, 6.2) | 4.3 (3.6, 5.3) |
| Current Smoker *** | 27.7 (25.0, 30.6) | 7.6 (5.4, 10.5) | 17.1 (15.5, 19.0) |
| Salt added at the Table | 9.3 (6.9, 12.6) | 11.5 (8.8, 15.0) | 10.5 (8.5, 12.7) |

\*p<0.05; \*\*p<0.01; \*\*\*p<0.001 for male: female differences

Estimates for 24-hour urine sodium and potassium concentration and proportions with abnormal values are shown in Table 3. Mean sodium excretion was 3582 mg/day (males 3943 mg/day, females 3245 mg/day, p<0.001). Mean potassium excretion was 2052 mg/day (males 2210 mg/day, females 1905 mg/day, p=0.001). Overall, 67% of participants were classified as having high sodium consumption (urine sodium excretion ≥ 2000 mg/24 hours); this was higher in men compared to women (73% vs. 61%, p<0.001). Prevalence of low potassium consumption (< 3510 mg/24 hour) was 89% and was higher among women (92% vs 85%, p<0.001). Approximately 74% of participants had high sodium to potassium ratio (≥ 1.0), higher in men compared to women (76% vs. 71%). Prevalence of high sodium consumption and low potassium consumption by age categories for men, women and the total population are shown in Figures 1A and 1B. Estimates show high prevalence of high sodium and low potassium consumption in all age groups with older men and women having the lowest prevalence estimates.

**Table 3:** Mean Urine Sodium and Potassium Excretion1 and Estimated Prevalence of High salt Consumption, Low Potassium Consumption and High Sodium/Potassium Ratio.

| Characteristic | Male<br>n = 368 | Female<br>n = 641 | Total<br>N = 1009 | p-value for<br>Male: Female<br>differences |
| --- | --- | --- | --- | --- |
|  | Mean (95% CI) | Mean (95% CI) | Mean (95% CI) |  |
| Urine Sodium [PAHO] (mg/24hr) | 3943 (3741, 4145) | 3245(3059, 3431) | 3582 (3441, 3724) | <0.001 |
| Urine Potassium [PAHO] (mg/24hr) | 2210 (2105, 2314) | 1905 (1790, 2019) | 2052 (1976, 2129) | <0.001 |
| Sodium/Potassium Ratio (mg/24hr) | 2.13 (1.00, 2.30) | 2.08 (1.91, 2.25) | 2.10 (1.99, 2.22) | 0.68 |
|  | % (95% CI) | % (95% CI) | % (95% CI) |  |
| High Sodium Consumption ($\geq 2000$ mg/24 hrs) | 72.8 (68.5, 76.8) | 60.7 (57.3, 64.1) | 66.6 (63.9, 69.2) | <0.001 |
| Low Potassium Consumption (< 3510 mg/24 hrs) | 85.1 (82.2, 87.6) | 92.3 (89.6, 94.3) | 88.8 (87.0, 90.4) | <0.001 |
| High Sodium/Potassium Ratio ($\geq 1.0$) | 76.2 (72.8, 79.3) | 71.1 (68.2, 73.7) | 73.5 (71.2, 75.7) | 0.016 |

<sup>1</sup>Mean 24-hour urine sodium and potassium were computed using the Pan American Health Organization formula as defined in text.

**Figure 1A:**
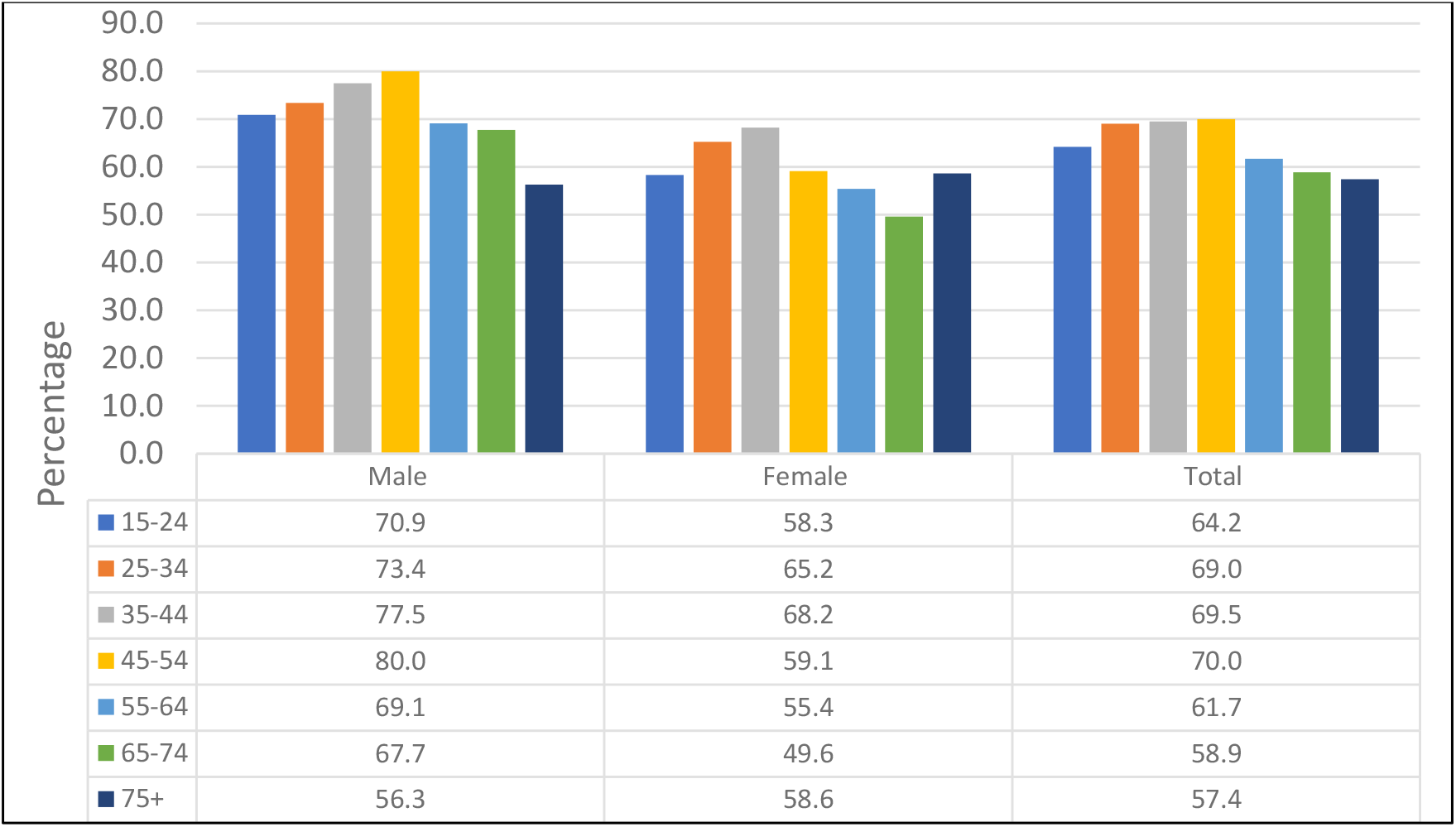
Prevalence of High Urine Sodium Excretion. P-value for association with age category: male, p=0.082, female p=0.219, total p= 0.019

**Figure 1B:**
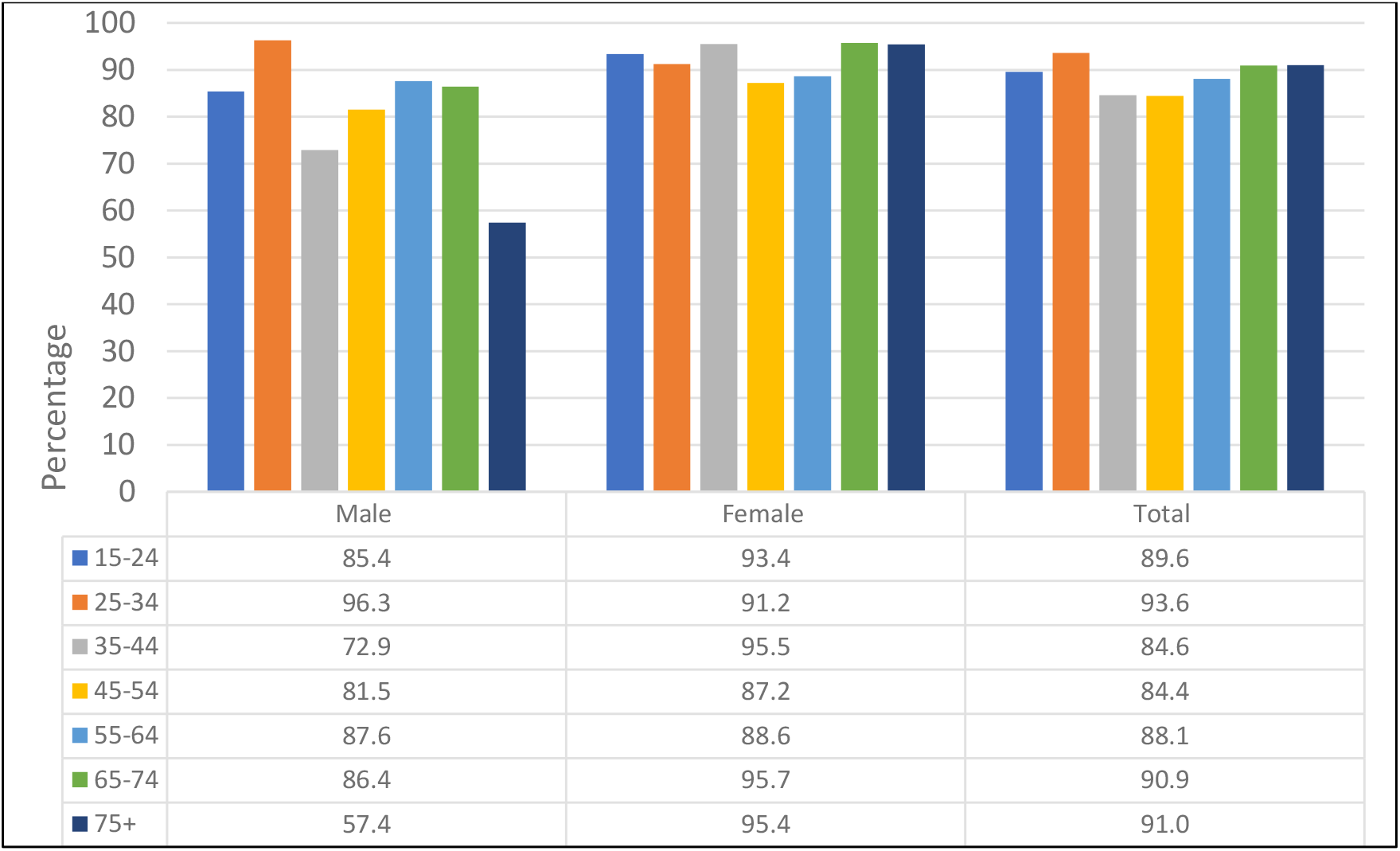
Prevalence of Low Urine Potassium Excretion. P-value for association with age category: male, p<0.001, female p=0.119, total p= 0.003

Participant characteristics based on sodium (high vs normal) and potassium consumption (low vs adequate) are presented in Tables S3 and S4 in the supplementary file. A higher proportion of men had high sodium consumption, while a higher proportion of women had normal sodium consumption (p<0.001). Persons who were cigarette smokers were more likely to have high sodium consumption, while those with low GFR were less likely to have high sodium consumption. Significant associations with low potassium consumption were seen for sex, blood pressure category, BMI categories, education level, high cholesterol, low GFR and current smoking.

Associations between sodium and potassium consumption and biomedical characteristics were also explored in bivariate AVOVA models using log-transformed 24-hour urine sodium and potassium. These findings are summarised in Tables S6 and S7 in the supplementary file. For sodium consumption, significant associations were seen for age (both continuous and categorical) for the total sample and for females. There were also significant associations for blood pressure categories, glucose categories, obesity, physical activity, low GFR and current smoking. There was evidence for significant sex interaction (p<0.05) for prehypertension and borderline significant interactions (p<0.1) for hypertension, high school education and current smoking. For potassium consumption, significant positive associations were seen for age, prehypertension, hypertension, diabetes, obesity, and high cholesterol. Inverse associations were seen for prehypertension and for high school education among males only. Significant sex interactions were seen for prehypertension, impaired fasting glucose, overweight status, and high school education. Given the significant sex-interactions identified in the bivariate analysis, we report sex-specific multivariable models.

Table 4 shows the results of sex-specific multivariable ANOVA models evaluating the association between sodium consumption and covariates including age category, blood pressure category, blood glucose level, BMI category, low GFR, education level, household possession category, current smoking and physical activity levels using log-transformed 24-hour urine sodium as the outcome variable. We report β-coefficients with 95% confidence intervals (95%CI) as well as the percent difference in 24-hour urine sodium compared to the reference category – in this case the youngest age group category. The percent difference was calculated as the (exponential of the β-coefficient – 1) x 100). Among males, older age was associated with lower urine sodium; the age group 55-74 years was associated with a 22% decrease in 24-hour urine sodium compared to age 15-34 years (p=0.049) and being 75 years and older was associated with a 29% decrease (p=0.032). Being in the prehypertension blood pressure category was associated with a 14% lower urine sodium compared to persons with normal blood pressure. Compared with normal BMI, obesity was associated with a 35% increase in urine sodium excretion (p=0.003), while current smoking was associated with a 25% increase in urine sodium excretion compared to non-smoking (p=0.008). There were no significant associations with household possession category, education level, or physical activity level for the male-specific model. Among females, hypertension and impaired fasting glucose were associated with 20% and 18% decreases in urine sodium respectively (p=0.016 [hypertension], p=0.001 [IFG]). Obesity was associated with a 57% increase in salt consumption compared to normal BMI (p<0.001), while high physical activity levels were associated with a 13% decrease in urine sodium (p=0.039). Having a low GFR was associated with a 32% decrease in urine sodium excretion. Smoking and household possession were excluded from the analysis for the female-specific model, given the criteria used for inclusion in the final multivariable models.

**Table 4:**
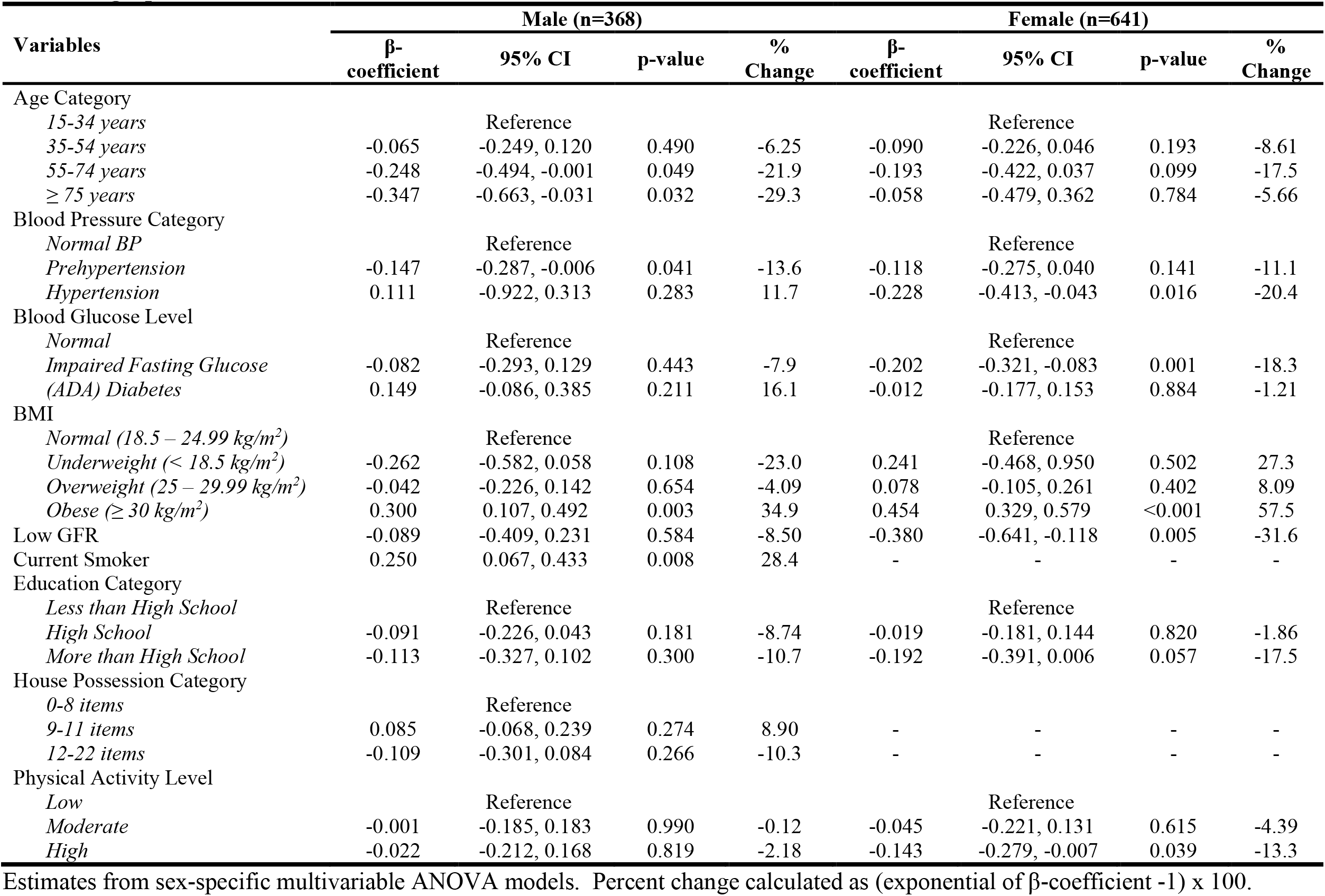
Sex-specific Multivariable ANOVA Models for the association between 24-hour urine sodium (log transformed) and sociodemographic, biomedical, and clinical characteristics.

Table 5 shows the results of the sex-specific multivariable ANOVA model for 24-hour log-transformed urinary potassium. Among men, the presence of pre-hypertension was associated with a 20% decrease in urinary potassium (p=0.004), while impaired fasting glucose, diabetes and obesity were associated with higher potassium (15% for IFG, p=0.026, 23% for diabetes, p=0.015; 43% increase with obesity, p<0.001). Secondary education was also found to be associated with a 17% decrease in urinary potassium in males. Among women, prehypertension was associated with a 15% increase in urinary potassium (p=0.034). Overweight, obesity and high cholesterol were also associated with higher potassium. Overweight was associated with a 28% increase (p=0.001), obesity with a 58% increase (p<0.001) and high cholesterol with a 21% increase (p<0.001).

**Table 5:** Sex-specific Multivariable ANOVA Models for the association between 24-hour urine potassium (log transformed) and sociodemographic, biomedical, and clinical characteristics.

| Variables | Male (n=368) |  |  |  | Female (n=641) |  |  |  |
| --- | --- | --- | --- | --- | --- | --- | --- | --- |
| | β-coefficient | 95% CI | p-value | % Change | β-coefficient | 95% CI | p-value | % Change |
| Age Category |  |  |  |  |  |  |  |  |
| 15-34 years | Reference |  |  |  | Reference |  |  |  |
| 35-54 years | 0.059 | -0.059, 0.177 | 0.323 | 6.08 | 0.113 | 0.010, 0.216 | 0.032 | 12.0 |
| 55-74 years | 0.001 | -0.141, 0.142 | 0.995 | 0.05 | 0.634 | -0.022, 0.148 | 0.142 | 6.55 |
| $\geq 75$ years | -0.116 | -0.345, 0.112 | 0.315 | -11.0 | 0.034 | -0.136, 0.204 | 0.692 | 3.47 |
| Blood Pressure Category |  |  |  |  |  |  |  |  |
| Normal BP | Reference |  |  |  | Reference |  |  |  |
| Prehypertension | -0.219 | -0.366, -0.072 | 0.004 | -19.7 | 0.137 | 0.005, 0.270 | 0.043 | 14.7 |
| Hypertension | -0.013 | -0.170, 0.144 | 0.868 | -1.31 | 0.053 | -0.059, 0.165 | 0.350 | 5.45 |
| Blood Glucose Level |  |  |  |  |  |  |  |  |
| Normal | Reference |  |  |  | Reference |  |  |  |
| Impaired Fasting Glucose | 0.140 | 0.017, 0.262 | 0.026 | 15.0 | -0.091 | -0.210, 0.421 | 0.085 | -8.7 |
| (ADA) Diabetes | 0.209 | 0.042, 0.376 | 0.015 | 23.2 | -0.004 | -0.095, 0.087 | 0.932 | -0.39 |
| BMI |  |  |  |  |  |  |  |  |
| Normal (18.5 – 24.99 kg/m<sup>2</sup>) | Reference |  |  |  | Reference |  |  |  |
| Underweight (< 18.5 kg/m<sup>2</sup>) | -0.154 | -0.378, 0.070 | 0.177 | -14.2 | 0.106 | -0.210, 0.421 | 0.508 | 11.2 |
| Overweight (25 – 29.99 kg/m<sup>2</sup>) | -0.004 | -0.115, 0.106 | 0.939 | -0.43 | 0.250 | 0.106, 0.395 | 0.001 | 28.4 |
| Obese ($\geq 30$ kg/m<sup>2</sup>) | 0.357 | 0.201, 0.513 | <0.001 | 42.9 | 0.455 | 0.330, 0.580 | <0.001 | 57.6 |
| High Cholesterol | 0.090 | -0.031, 0.210 | 0.144 | 9.38 | 0.188 | 0.089, 0.287 | <0.001 | 20.7 |
| Education Category |  |  |  |  |  |  |  |  |
| Less than High School | Reference |  |  |  | Reference |  |  |  |
| High School | -0.187 | -0.285, -0.089 | <0.001 | -17.0 | 0.020 | -0.071, 0.110 | 0.666 | 1.99 |
| More than High School | -0.957 | -0.264, 0.072 | 0.261 | -9.13 | 0.123 | -0.001, 0.247 | 0.052 | 13.1 |

Estimates from sex-specific multivariable ANOVA models. Percent change calculated as (exponential of β-coefficient -1) x 100.

As a form of sensitivity analysis, prevalence ratios were also estimated using Poisson regression for the covariates and outcomes described in the multivariable ANOVA models above. These are shown in Tables S8 and S9 in the supplementary file. The findings in these models were generally similar to those in the ANOVA models. Men older than 75 years were found to have a lower prevalence of high sodium consumption (PR 0.74, p=0.029), while there was no significant association with age for women. Current smoking was associated with a higher prevalence of high sodium (PR 1.19, p=0.038) in men, and this variable was not analysed for the female subgroup. Women with prehypertension and hypertension had lower prevalence of high sodium consumption (PR 0.75 [prehypertension], PR 0.74 [hypertension], p>0.001 for both). Among women, impaired fasting glucose was also associated with a lower prevalence of high sodium consumption (PR 0.80, p<0.001), while obesity was associated with higher prevalence of high sodium (PR 1.47, p<0.001). For low potassium consumption (Table S9), among men, older age, pre-hypertension, and secondary education were associated with a higher prevalence of low potassium. Men older than 75 years and men with prehypertension were half as likely to have low potassium consumption, (PR for age > 75 years 0.49 (p=0.034) and PR for pre-hypertension 0.51 (p=0.012)). On the other hand, men with obesity had an almost 2-fold increase in the prevalence of low potassium consumption (PR 1.99, p<0.001). Among women, obesity was associated with a 4-fold increase in the prevalence of low potassium consumption (PR 4.50, p<0.001), and overweight with an almost 3-fold increase (PR 2.817, p=0.005). High cholesterol was associated with a 2-fold increase in the prevalence of low potassium (PR 2.09, p=0.022) among women.

To assess whether the models using multiple imputation would yield similar inferences to models without imputation we re-ran the ANOVA models using complete case analyses. These models included 208 males and 510 females, compared to 368 and 641 in the full models with imputation. Findings for the complete case analyses are shown in Tables S10 and S11. The findings in the complete cases analyses were generally similar in direction and magnitude; we noted that underweight among males was now statistically significant, while among women, age category 55-74 years and education categories were now significant. For the potassium, complete cases analyses findings were again similar to those in the multiple imputation analyses except that prehypertension was significantly associated with among females.

## DISCUSSION

In this study we found that estimated mean sodium consumption is high among Jamaican adults while mean potassium consumption is quite low. Correspondingly, two thirds of Jamaicans are classified as having high sodium consumption and almost 90% have low potassium consumption when assessed by recommended cut-points from the WHO. Sodium consumption was directly associated with obesity in both men and women, but associations differed by sex for other factors. Potassium consumption was also significantly associated with obesity in both men and women, but again, except for an inverse associate for prehypertension in both men and women, other associations for potassium differed by sex.

Estimated mean sodium consumption in this study was higher than that reported among persons 25-64 years old in Barbados (2656 mg/day) but was lower than the estimated global mean (3950 mg/day), and similar to estimates from the United States of America [USA] (3608 mg/day) (4, 39, 46). The estimates in this study were also similar to that reported from the Spanish Town Cohort in the 1990s (23), suggesting a pattern of high sodium and low potassium consumption for almost three decades. As seen in our study, mean sodium consumption estimates were higher among men in Barbados, the USA and in global estimates. Mean estimates in the USA were similar among black and white race categories. Estimated prevalence of high sodium consumption was similar in Jamaica and Barbados, 67% in both, but lower than in the USA (75%), even though the USA cut point was 2300 mg/day, while the WHO cut point of 2000 mg/day was used for Jamaica and Barbados. Estimated mean potassium consumption was higher in Jamaica than in Barbados (1469 mg/day) and was again similar to that reported in the USA (2155 mg/day) (39, 46). The findings in Jamaica were also similar to reports from a recent systematic review looking at sodium and potassium excretion in the Americas, where mean sodium excretion was 157 mmol/day (3617 mg/day) and mean potassium excretion of 58 mmol/day (2256 mg/day) (47). Sodium consumption in our study was however lower than reports from Moldova and Northern China with estimates of 3972 mg/day and 5336 mg/day (48, 49). In these latter two studies, potassium excretion was 2842 mg/day and 1595 mg/day in Moldova and Northern China, respectively.

The analytic models in this study were primarily exploratory as we sought to identify factors associated with sodium and potassium consumption, particularly factors which could be targeted in future public health interventions. The strongest association found was for obesity which was associated with a 35% to 58% higher sodium and potassium consumption and was significant in all the models. Older age was associated with lower sodium consumption in men only, while reduced GFR was associated with lower sodium in women only, and cigarette smoking associated with higher sodium in men only. The association between obesity and sodium and potassium intake is noteworthy. Similar findings have been reported from studies in the United States (USA), United Kingdom (UK) and Korea (50-53). In the USA each 1 g increase in daily sodium consumption was associated with 3.8 units higher BMI, while in the UK a 1 g per day increase in sodium consumption was associated with a 20% increase in the risk of obesity (50, 52). While an association between high salt intake and increase caloric consumption may be a possible mechanism contributing to these effects, the studies cited above assessed the effect of caloric intake and found the association was independent of caloric intake. It has been hypothesized that salt intake may have direct effects on appetite through stimulation of ghrelin (54) or may even have addictive properties through effects on opiate and dopamine receptors, resulting in overeating associated with ‘salt withdrawal’ (55). These findings suggest that high salt consumption may be a contributor to the increasing prevalence of obesity in Jamaica, further supporting the need for salt reduction interventions.

In this study sodium consumption was inversely associated with hypertension among women but was not associated with hypertension among men. There was no significant association between hypertension and potassium intake. These findings differed from the expected finding of a direct association with sodium intake and inverse association with potassium intake reported in other studies (56, 57). The inverse association among women may be due to reverse causality, where women with hypertension have been advised to reduce salt intake. That the estimates in the current study were based on single spot urine sodium samples may also introduce greater variability in measurements thus reducing the ability to show an effect. The finding of inverse associations with prehypertension and sodium consumption among men and inverse associations between prehypertension and potassium consumption in both men and women requires further exploration in future studies.

We found age category was inversely associated with sodium consumption but was statistically significant only for men in the 55-74 and 75 and older age groups. The study from Barbados (39) did not report a significant association between age and sodium consumption in multivariable models, although sex specific analyses were not reported. In the USA study, bivariate analyses suggested similar sodium consumption in age categories and higher potassium consumption among older individuals (46). Similar to our findings there was no association with education level in Barbados or in the USA. There was also no association between sodium consumption and physical activity level in the USA. Men who reported current smoking had a 28% higher sodium consumption compared to non-smokers. Neither the USA nor the Barbadian study reported associations between sodium consumption and cigarette smoking. A preliminary Korean study reported that current smokers had higher mean sodium intake and preference for salty foods (58). We also noted that there was an inverse association between impaired fasting glucose and sodium consumption among women and a direct association with potassium intake for both impaired fasting glucose and diabetes among men. None of the studies reviewed reported specific associations between salt intake and diabetes or impaired fasting glucose, although diabetes was often included as a covariate in multivariable models. These findings should therefore be explored in future studies. With regards to the finding of an inverse association between low GFR and sodium intake, this may also be related to reverse causality resulting from reduced sodium consumption in patients known to have CKD or conditions that place persons at risk of CKD as well as sodium retention associated with CKD itself. Further studies would be required to better understand this association.

This study has some limitations. Firstly, sodium and potassium levels were measured using spot urine samples, rather than the preferred 24-hour urine samples. However, recent studies have shown that spot urine sample estimates can give good indications of mean population salt intake and the need for population-based action (59). The consistency of our findings with those from studies based on 24-hour urine collections also adds to the plausibility of our estimates. Secondly, spot urine samples were only available for a subset of the survey participants and some participants had missing data. We used multiple imputation to fill in missing data, thus minimising potential bias and reduced power.

Strengths of this study include the fact that it was population based and used survey weights to ensure that estimates are nationally representative. These data represent the first report of national estimates of sodium consumption. Other strengths include the use of standardized measurement procedures to ensure data quality and the use of multiple imputation to account for missing data.

These findings have important public health implications for Jamaica. Given the high prevalence of hypertension and prehypertension, major contribution of cardiovascular disease to morbidity and mortality, and the more recent findings of high prevalence of chronic kidney disease (20, 22, 60-62), there is an urgent need to increase awareness of the dangers of high salt diets and the implementation of public health interventions that take a population-based approach to reduce sodium intake and increase potassium intake. The high prevalence, even in the younger age group, suggests that intervention should start early and should include strategies to engage younger persons. Further research should identify sources of sodium and potassium in the diet, evaluate strategies to implement interventions, and their effectiveness in the Jamaican context.

## Data Availability

All data produced in the present study are available upon reasonable request to the authors

## ACKNOWLEDGEMENTS

The authors would like to thank the Ministry of Health and Wellness and the National Health Fund Jamaica, which provided funding for the Jamaica Health and Lifestyle Survey III in 2016/17 and the Jamaica Salt Consumption, Knowledge, Attitudes and Practice (Salt-KAP) Study. We would also like to thank the JHLS-III field staff, other JHLS-III collaborators, JHLS-III participants and the CAIHR/ERU administrative staff and the unit driver for their support.

## AUTHOR CINTRIBUTIONS

**TSF**: conceptualized paper, developed data-analysis plan along with NYC, interpreted data, wrote first draft of manuscript, and revised the manuscript after critical reviewed by co-authors.

**NYC:** contributed to data collection; lead data preparation, lead the data analysis, contributed to interpretation of data, and critically reviewed the manuscript.

**KWK:** contributed to data collection, contributed to interpretation of data and critical review of the manuscript.

**MKTR:** contributed to data collection, contributed to interpretation of data and critical review of the manuscript.

**NRB:** contributed to data collection, contributed to interpretation of data and critical review of the manuscript.

**TD:** contributed to data collection, contributed to study conceptualization, contributed to interpretation of data and critical review of the manuscript.

**AG:** contributed to data collection, contributed to interpretation of data and critical review of the manuscript.

**KMGJ:** contributed to study conceptualization, contributed to interpretation of data and critical review of the manuscript.

**IG:** contributed to data collection, contributed to interpretation of data and critical review of the manuscript.

**SSW:** contributed to data collection, contributed to interpretation of data and critical review of the manuscript.

**JAM:** contributed to interpretation of data and critical review of the manuscript.

**EW:** contributed to interpretation of data and critical review of the manuscript.

**CACM:** contributed to data collection, contributed to interpretation of data and critical review of the manuscript.

**SGA:** contributed to data analysis plan, contributed to interpretation of data and critical review of the manuscript.

**ALB:** contributed to interpretation of data and critical review of the manuscript.

**JH:** contributed to preparation of first draft of manuscript; contributed to interpretation of data and critical review of the manuscript.

**RB:** contributed to preparation of first draft of manuscript; contributed to interpretation of data and critical review of the manuscript.

**SEE:** contributed to data collection, contributed to interpretation of data and critical review of the manuscript.

**SRM:** contributed to data collection, contributed to interpretation of data and critical review of the manuscript.

**SS:** contributed to interpretation of data and critical review of the manuscript.

**RJW:** contributed to data collection, contributed to interpretation of data and critical review of the manuscript.

## Notes

**Previous presentations** - Conference presentation: Caribbean Public Health Agency 66th Annual Scientific Meeting 2022. Kingston, Jamaica. Abstract in West Indian Medical Journal at https://www.mona.uwi.edu/fms/wimj/system/files/article_pdfs/carpha_2022_oral_abstracts.pdf

**Funding Support** - This project was supported by a research grant from the National Health Fund (Jamaica) – Grant # HSF 597 - The Jamaica Health and Lifestyle Survey 2016-2017 was supported by the National Health Fund (Jamaica) – Grant # HPP 315 No funding was received from the National Institutes of Health, Wellcome Trust or Howard Hughes Medical Institute

**Conflicts of Interest** The authors declare that there are no relevant conflicts of interest.

### Competing Interest Statement

The authors have declared no competing interest.

### Clinical Protocols

https://doi.org/10.12688/f1000research.122619.1

### Funding Statement

This project was supported by a research grant from the National Health Fund (Jamaica). Grant # HSF 597.
The Jamaica Health and Lifestyle Survey 2016-2017 was supported by the National Health Fund (Jamaica). Grant # HPP 315

### Author Declarations

The Ethics Committees of the University of the West Indies Mona Faculty of Medical Sciences and the Ministry of Health and Wellness, Jammaica gave ethical approval for this work.

