## Supplementary Tables for "Sodium and Potassium Consumption in Jamaica: National Estimates and Associated Factors from the Jamaica Health and Lifestyle Survey 2016-2017"

---

Trevor S FERGUSON[a], Novie OM YOUNGER-COLEMAN[a], Karen WEBSTER-KERR[b], Marshall K. TULLOCH-REID[a], Nadia R BENNETT[a], Tamu DAVIDSON[b], Andriene S GRANT[b], Kelly-Ann M. GORDON-JOHNSON[c], Ishtar GOVIA[a], Suzanne SOARES-WYNTER[d], Joette A MCKENZIE[a], Evelyn WALKER[a], Colette A CUNNINGHAM-MYRIE[e], Simon G ANDERSON[g], Alphanso L BLAKE[a], James HO[a], Robyn STEPHENSON[g], Sharmaine E EDWARDS[b], Shelly R MCFARLANE[e], Simone SPENCE[b], Rainford J WILKS[a]

[a]*Epidemiology Research Unit, Caribbean Institute for Health Research Institute, The University of the West Indies, Mona, Kingston, Jamaica.*

[b]*Ministry of Health and Wellness, Kingston, Jamaica.*

[c]*Caribbean Regional Office, Centers for Disease Control and Prevention, United States Embassy, Jamaica*

[d]*Tropical Metabolism Research Unit, Caribbean Institute for Health Research Institute, The University of the West Indies, Mona, Kingston, Jamaica.*

[e]*Department of Community Health and Psychiatry, Faculty of Medical Sciences, The University of the West Indies, Mona, Kingston, Jamaica.*

[f]*George Alleyne Chronic Disease Research Centre, Caribbean Institute for Health Research Institute, The University of the West Indies, Cave Hill, Bridgetown, Barbados.*

[g]*Western Regional Health Authority, Hanover, Jamaica*

**Corresponding author**
Professor Trevor S. Ferguson
Epidemiology Research Unit
Caribbean Institute for Health Research
University of the West Indies, Mona
Kingston 7, Jamaica


**Table S1: Items Included in Household Assets List**

| Included Household Assets | |
|---|---|
| 1. | Gas stove/Electric Stove |
| 2. | Refrigerator or freezer |
| 3. | Microwave oven |
| 4. | Air conditioner |
| 5. | Fan |
| 6. | Telephone (land line or cell) |
| 7. | Radio/cassette player /Stereo equipment/component set |
| 8. | Electronic Gaming Equipment |
| 9. | Video cassette recorder/ DVD |
| 10. | Washing machine |
| 11. | Clothes Dryer |
| 12. | TV sets |
| 13. | Cable TV |
| 14. | Water Heater (Solar or Electric) |
| 15. | Water Tank |
| 16. | Bicycle |
| 17. | Motorbike |
| 18. | Car or another vehicle |
| 19. | Computer/ Tablet |
| 20. | Computer Accessories /Printer/Fax/Scanner |
| 21. | Smart Phone |
| 22. | Internet service |

**Table S2: Number and Proportion of Missing values for variables included in the analyses**

| Variable | Number of missing | Number of non-missing values | % Missing |
|---|---|---|---|
| Height | 19 | 990 | 1.9 |
| Weight | | | |
| Body Mass Index | 28 | 981 | 2.8 |
| Systolic Blood Pressure | 10 | 999 | 1.0 |
| Diastolic Blood Pressure | 10 | 999 | 1.0 |
| Fasting glucose | 60 | 949 | 5.9 |
| Fasting cholesterol | 84 | 925 | 8.3 |
| Healthy diet | | | |
| Current non-smoker | 347 | 662 | 34.4 |
| Physical Activity | 39 | 970 | 3.9 |
| Education | 32 | 977 | 3.2 |
| Household Assets | 2 | 1007 | 0.2 |
| Median Land Value | | | |

**Table S3: Survey weighted means for participant characteristics for males, females and total sample.**

| Characteristic | Male n = 368 Mean (95% CI) | Female n = 641 Mean (95% CI) | Total N = 1009 Mean (95% CI) | p-value for Male: Female differences |
|---|---|---|---|---|
| Age (years) | 40.5 (40.4, 40.6) | 39.7 (39.6, 39.9) | 40.1 (40.0, 40.2) | <0.001 |
| Weight (kg) | 79.2 (76.9, 81.5) | 78.1 (76.4, 79.7) | 78.6 (77.3, 79.9) | 0.469 |
| Height (cm) | 173.0 (172.2, 173.7) | 162.9 (161.6, 164.1) | 167.7 (167.0, 168.4) | <0.001 |
| Body mass index ($kg/m^2$) | 25.9 (25.1, 26.7) | 29.5 (28.9, 30.1) | 27.8 (27.3, 28.3) | <0.001 |
| Systolic Blood Pressure (mmHg) | 128.9 (127.7, 130.2) | 124.3 (122.9, 125.7) | 126.6 (125.5, 127.6) | <0.001 |
| Diastolic Blood Pressure (mmHg) | 80.0 (79.2, 80.9) | 81.2 (80.1, 82.3) | 80.7 (79.9, 81.4) | 0.078 |
| Fasting glucose (mmol/l) | 5.61 (5.53, 5.69) | 5.90 (5.69, 6.11) | 5.76 (5.65, 5.87) | 0.016 |
| Total Cholesterol (mmol/l) | 4.42 (4.33, 4.51) | 4.45 (4.38, 4.52) | 4.43 (4.37, 4.50) | 0.619 |
| Estimated GFR ($ml/min/1.73m^2$) | 102.0 (100.3, 103.6) | 105.1 (102.9, 107.4) | 103.6 (102.0, 105.2) | 0.010 |
| Spot urine sodium (mg/l) | 3341.7 (3189.7, 3493.8) | 2971.3 (2866.2, 3076.4) | 3150.5 (3059.4, 3241.5) | <0.001 |
| Spot urine potassium (mg/l) | 2127.2 (2021.2, 2233.3) | 2111.8 (2018.1, 2205.5) | 2119.3 (2047.0, 2191.5) | 0.825 |
| 24-hour urine creatinine (CKD-EPI, mg) | 1653.1 (1624.5, 1681.7) | 1264.4 (1243.6, 1285.2) | 1452.4 (1436.6, 1468.2) | <0.001 |

**Table S4: Proportion (%) of Participant Characteristics By Salt Consumption Categories**

| Characteristic | | Normal Salt Consumption <2000 mg/day % (95% CI) | High Salt Consumption ≥2000 mg/day % (95% CI) |
|---|---|---|---|
| Sex*** | | | |
| | Male | 39.3 (35.1, 43.6) | 52.9 (50.9, 54.9) |
| | Female | 60.7 (56.4, 64.9) | 47.1 (45.1, 49.1) |
| Blood Pressure Category | | | |
| | Normal BP | 26.8 (22.0, 32.3) | 32.7 (29.8, 35.8) |
| | Prehypertension | 39.1 (34.1, 44.4) | 36.8 (33.6, 40.1) |
| | Hypertension | 34.1 (31.0, 37.3) | 30.5 (27.8, 33.4) |
| Blood Glucose Level | | | |
| | Normal | 55.2 (49.9, 60.3) | 58.4 (54.7, 62.1) |
| | Impaired Fasting Glucose | 33.6 (28.5, 39.1) | 28.3 (25.0, 31.9) |
| | (ADA) Diabetes | 11.3 (8.7, 14.5) | 13.3 (10.9, 16.1) |
| BMI | | | |
| | Normal (18.5 – 24.99 $kg/m^2$) | 40.5 (35.9, 45.3) | 37.8 (34.6, 41.0) |
| | Underweight (< 18.5 $kg/m^2$) | 6.9 (4.2, 11.2) | 3.2 (2.0, 4.9) |
| | Overweight (25 – 29.99 $kg/m^2$) | 25.9 (21.4, 30.9) | 26.6 (23.7, 29.8) |
| | Obese (≥ 30 $kg/m^2$) | 26.8 (22.3, 31.8) | 32.5 (30.0, 35.0) |
| Physical Activity Level | | | |
| | Low | 33.2 (28.1, 38.7) | 35.5 (32.8, 38.3) |
| | Moderate | 29.3 (25.0, 34.1) | 26.8 (23.2, 30.8) |
| | High | 37.5 (33.3, 41.8) | 37.7 (33.9, 41.6) |
| Education | | | |
| | Less than High School | 29.0 (25.8, 32.3) | 24.5 (22.3, 26.9) |
| | High School | 54.6 (49.6, 59.5) | 60.5 (56.7, 64.1) |
| | More than High School | 16.4 (12.1, 22.0) | 15.0 (12.1, 18.5) |
| Household Possessions | | | |
| | ≤8 items | 35.7 (30.5, 41.2) | 33.0 (30.5, 35.6) |
| | 9-11 items | 30.5 (26.4, 34.9) | 34.7 (30.8, 38.8) |
| | 12-22 items | 33.9 (29.1, 39.0) | 32.3 (28.2, 36.8) |
| High Cholesterol | | 17.3 (14.3, 20.8) | 21.4 (18.7, 24.4) |
| Low GFR** | | 5.9 (4.4, 7.7) | 3.5 (2.7, 4.6) |
| Current Smoker*** | | 12.0 (9.3, 15.5) | 19.8 (17.8, 22.0) |
| Added Salt at the Table | | 9.3 (6.7, 12.8) | 11.0 (8.7, 13.8) |

*p<0.05; **p<0.01; ***p<0.001; Proportions are survey weighted using Stata's '*svy: prop'* command. P-values are for overall difference in proportions across all categories from Pearson's chi squared test using Stata's '*svy: tab'* command.

**Table S5: Proportion (%) of Participant Characteristics By Potassium Consumption Categories**

| Characteristic | | Adequate Potassium Consumption (≥3510 mg/day) % (95% CI) | Low Potassium Consumption (<3510 mg/day) % (95% CI) |
|---|---|---|---|
| Sex*** | | | |
| | Male | 64.0 (55.5, 71.6) | 46.4 (45.3, 47.4) |
| | Female | 36.0 (28.4, 44.5) | 53.6 (52.6, 54.7) |
| Blood Pressure Category* | | | |
| | Normal BP | 24.6 (19.5, 30.5) | 31.6 (29.0, 34.3) |
| | Prehypertension | 36.1 (29.8, 43.0) | 37.6 (34.6, 40.6) |
| | Hypertension | 39.3 (33.7, 45.2) | 30.9 (28.5, 33.4) |
| Blood Glucose Level | | | |
| | Normal | 52.3 (44.4, 60.1) | 58.2 (55.2, 61.1) |
| | Impaired Fasting Glucose | 35.2 (27.9, 43.2) | 29.1 (26.5, 31.9) |
| | (ADA) Diabetes | 12.6 (8.8, 17.6) | 12.7 (10.7, 15.0) |
| BMI*** | | | |
| | Normal (18.5 – 24.99 kg/m$^2$) | 28.2 (22.4, 34.8) | 40.3 (37.6, 43.1) |
| | Underweight (< 18.5 kg/m$^2$) | 3.2 (0.9, 10.2) | 4.6 (3.6, 6.0) |
| | Overweight (25 – 29.99 kg/m$^2$) | 19.3 (14.7, 24.8) | 26.8 (24.6, 29.0) |
| | Obese (≥ 30 kg/m$^2$) | 49.4 (41.7, 57.1) | 28.3 (25.9, 30.7) |
| Physical Activity Level | | | |
| | Low | 30.0 (23.5, 37.4) | 35.5 (32.5, 38.7) |
| | Moderate | 24.5 (17.7, 32.8) | 28.2 (25.0, 31.6) |
| | High | 45.5 (39.7, 51.5) | 36.3 (33.2, 39.5) |
| Education*** | | | |
| | Less than High School | 38.1 (31.4, 0.453) | 24.4 (22.5, 26.4) |
| | High School | 41.5 (33.9, 49.5) | 60.7 (57.3, 63.9) |
| | More than High School | 20.4 (16.4, 25.1) | 15.0 (12.1, 18.4) |
| Household Possessions | | | |
| | ≤8 items | 37.8 (31.3, 44.7) | 33.4 (30.6, 36.4) |
| | 9-11 items | 33.5 (27.1, 40.7) | 33.1 (30.3, 36.0) |
| | 12-22 items | 28.7 (23.4, 34.6) | 33.5 (30.1, 37.1) |
| High Cholesterol*** | | 31.4 (25.7, 37.7) | 18.7 (16.7, 20.9) |
| Low GFR** | | 17.1 (0.9, 3.2) | 4.6 (3.8, 5.7) |
| Current Smoker* | | 12.4 (9.1, 16.8) | 17.8 (16.0, 19.7) |
| Added Salt at the Table | | 11.0 (7.5, 15.7) | 10.6 (8.4, 13.3) |

*p<0.05; **p<0.01; ***p<0.001. Proportions are survey weighted using Stata's '*svy: prop*' command. P-values are for overall difference in proportions across all categories from Pearson's chi squared test using Stata's '*svy: tab*' command.

**Table S6: Bivariate models for association between 24-hour urine sodium (log transformed) and sociodemographic, biomedical, and clinical characteristics.**

| Characteristic | Male & Female combined | Male n = 368 | Female n = 641 | p-value for sex interaction |
|---|---|---|---|---|
| Age at last birthday (years) | -0.003 (-0.006, -0.001)** | -0.002 (-0.005, 0.001) | -0.005 (-0.008, -0.002)** | 0.201 |
| Age category | | | | |
| *15-34* | Reference | Reference | Reference | |
| *35-54* | -0.024 (-0.010, 0.052) | -0.015 (-0.132, 0.102) | -0.040 (-0.153, 0.073) | 0.780 |
| *55-74* | -0.177 (-0.281, -0.074)** | -0.105 (-0.251, 0.040) | -0.249 (-0.392, -0.106)** | 0.164 |
| *≥75* | -0.145 (-0.380, 0.091) | -0.112 (-0.403, 0.179) | -0.209 (-0.558, 0.140) | 0.657 |
| Blood Pressure Category | | | | |
| *Normal BP* | Reference | Reference | Reference | |
| *Prehypertension* | -0.072 (-0.191, 0.046) | -0.213 (-0.374, -0.058)** | 0.003 (-0.150, 0.156) | 0.027* |
| *Hypertension* | -0.074 (-0.192, 0.044) | 0.012 (-0.155, 0.179) | -0.162 (-0.298, -0.253)* | 0.060 |
| Blood Glucose Level | | | | |
| *Normal* | Reference | Reference | Reference | |
| *Impaired Fasting Glucose* | -0.149 (-0.265, -0.032)* | -0.130 (-0.319, 0.058) | -0.160 (-0.290, -0.030)* | 0.789 |
| *(ADA) Diabetes* | -0.008 (-0.160, 0.144) | 0.057 (-0.191, 0.304) | -0.016 (-0.184, 0.152) | 0.619 |
| BMI | | | | |
| *Normal (18.5 – 24.99 kg/m$^2$)* | Reference | Reference | Reference | |
| *Underweight (< 18.5 kg/m$^2$)* | -0.104 (-0.463, 0.255) | -0.290 (-0.603, 0.023) | 0.221 (-0.573, 1.01) | 0.175 |
| *Overweight (25 – 29.99 kg/m$^2$)* | -0.056 (-0.174, 0.063) | -0.097 (-0.277, 0.084) | 0.301 (-0.135. 0.196) | 0.331 |
| *Obese (≥ 30 kg/m$^2$)* | 0.230 (0.136, 0.324)*** | 0.288 (0.062, 0.514)* | 0.351 (0.234, 0.468)*** | 0.652 |
| Physical Activity Level | | | | |
| *Low* | Reference | Reference | Reference | |

| Characteristic | Male & Female combined | Male n = 368 | Female n = 641 | p-value for sex interaction |
|---|---|---|---|---|
| *Moderate* | -0.048 (-0.161, 0.064) | -0.132 (-0.303, 0.039) | -0.037 (-0.199, 0.125) | 0.447 |
| *High* | -0.055 (-0.148, 0.037) | -0.164 (-0.029, -0.037)* | -0.126 (-0.261, 0.001) | 0.668 |
| Education | | | | |
| *Less than High School* | Reference | Reference | Reference | |
| *High School* | 0.043 (-0.035, 0.121) | -0.020 (-0.111, 0.071) | 0.131 (-0.004, 0.265) | 0.077 |
| *More than High School* | -0.172 (-0.315, -0.029)* | -0.163 (-0.381, 0.055) | -0.105 (-0.279, 0.069) | 0.663 |
| Household Possessions | | | | |
| *≤8 items* | Reference | Reference | Reference | |
| *9-11 items* | 0.0612 (-0.065, 0.189) | 0.126 (-0.060, 0.312) | 0.0162 (-0.142, 0.175) | 0.314 |
| *12-22 items* | -0.078 (-0.196, 0.041) | -0.120 (-0.299, 0.060) | -0.027 (-0.172, 0.118) | 0.405 |
| High Cholesterol | 0.0197 (-0.091, 0.130) | 0.006 (-0.156, 0.169) | 0.052 (-0.098, 0.203) | 0.684 |
| Low GFR | -0.340 (-0.521, -0.158)*** | -0.193 (-0.442, 0.055) | -0.422 (-0.660, -0.185)** | 0.194 |
| Current Smoker | 0.267 (0.137, 0.396)*** | 0.286 (0.127, 0.446)** | -0.027 (-0.343, 0.289) | 0.080 |
| Salt added at the Table | -0.027 (-0.180, 0.127) | -0.042 (-0.192, 0.107) | 0.012 (-0.197, 0.220) | 0.624 |

*p<0.05; **p<0.01; ***p<0.001. Estimates derived from bivariate ANOVA models using survey weights and multiple imputation for missing data

**Table S7: Bivariate models for association between 24-hour potassium (log transformed) and sociodemographic, biomedical, and clinical characteristics.**

| Characteristic | Male & Female combined | Male n = 368 % (95% CI) | Female n = 641 % (95% CI) | p-value for sex interaction |
|---|---|---|---|---|
| Age at last birthday (years) | 0.005 (0.003, 0.007)*** | 0.004 (0.002, 0.006)*** | 0.005 (0.002, 0.008)*** | 0.529 |
| Age category | | | | |
| 15-34 | Reference | Reference | Reference | |
| 35-54 | 0.219 (0.142, 0.295)*** | 0.161 (0.052, 0.271)* | 0.267 (0.149, 0.385)*** | 0.221 |
| 55-74 | 0.213 (0.138, 0.289)*** | 0.200 (0.107, 0.294)*** | 0.222 (0.114, 0.330)*** | 0.758 |
| ≥75 | 0.178 (0.077, 0.279)** | 0.205 (0.081, 0.329)** | 0.124 (-0.420, 0.291) | 0.451 |
| Blood Pressure Category | | | | |
| Normal BP | Reference | Reference | Reference | |
| Prehypertension | 0.071 (-0.028, 0.170) | -0.156 (-0.307, -0.006)* | 0.245 (0.098, 0.392)** | <0.001 |
| Hypertension | 0.212 (0.128, 0.297)*** | 0.136 (0.010, 0.263)* | 0.246 (0.121, 0.372)*** | 0.250 |
| Blood Glucose Level | | | | |
| Normal | Reference | Reference | Reference | |
| Impaired Fasting Glucose | 0.712 (-0.027, 0.171) | 0.169 (0.040, 0.297)* | -0.013 (-0.150, 0.123) | 0.046 |
| (ADA) Diabetes | 0.163 (0.065, 0.260)** | 0.228 (0.073, 0.382)** | 0.133 (0.015, 0.251)* | 0.333 |
| BMI | | | | |
| Normal (18.5 – 24.99 $kg/m^2$) | Reference | Reference | Reference | |
| Underweight (< 18.5 $kg/m^2$) | -0.055 (-0.277, 0.168) | -0.202 (-0.451, 0.047) | 0.124 (-0.205, 0.453) | 0.073 |
| Overweight (25 – 29.99 $kg/m^2$) | 0.117 (0.023, 0.212)* | -0.012 (-0.119, 0.095) | 0.300 (0.151, 0.450)*** | 0.001 |
| Obese (≥ 30 $kg/m^2$) | 0.357 (0.261, 0.453)*** | 0.365 (0.202, 0.528)*** | 0.498 (0.364, 0.631)*** | 0.209 |
| Physical Activity Level | | | | |

| Characteristic | Male & Female combined | Male<br>n = 368<br>% (95% CI) | Female<br>n = 641<br>% (95% CI) | p-value for sex interaction |
|---|---|---|---|---|
| Low | Reference | Reference | Reference | |
| Moderate | -0.015<br>(-0.143, 0.113) | 0.004<br>(-0.170, 0.178) | -0.040<br>(-0.200, 0.121) | 0.688 |
| High | 0.042<br>(-0.068, 0.152) | 0.089<br>(-0.050, 0.228) | -0.094<br>(-0.260, 0.072) | 0.083 |
| Education | | | | |
| Less than High School | Reference | Reference | Reference | |
| High School | -0.158<br>(-0.232, -0.084)*** | -0.228<br>(-0.320, -0.136)*** | -0.071<br>(-0.171, 0.029) | 0.019 |
| More than High School | -0.111<br>(-0.226, 0.003) | -0.082<br>(-0.283, 0.120) | -0.079<br>(-0.194, 0.036) | 0.982 |
| Household Possessions | | | | |
| ≤8 items | Reference | Reference | Reference | |
| 9-11 items | 0.035<br>(-0.084, 0.153) | 0.071<br>(-0.148, 0.289) | 0.008<br>(-0.124, 0.140) | 0.640 |
| 12-22 items | 0.021<br>(-0.095, 0.137) | 0.078<br>(-0.110, 0.266) | -0.026<br>(-0.122, 0.071) | 0.264 |
| High Cholesterol | 0.208<br>(0.126, 0.291)*** | 0.201<br>(0.61, 0.340)** | 0.228<br>(0.120, 0.336)*** | 0.766 |
| Low GFR | -0.063<br>(-0.176, 0.050) | -0.093<br>(-0.243, 0.058) | -0.025<br>(-0.182, 0.131) | 0.538 |
| Current Smoker | 0.000<br>(-0.129, 0.128) | -0.032<br>(-0.189, 0.125) | -0.112<br>(-0.325, 0.102) | 0.548 |
| Added Salt at the Table | -0.075<br>(-0.211, 0.061) | -0.089<br>(-0.313, 0.135) | -0.051<br>(-0.201, 0.098) | 0.768 |

*p<0.05; **p<0.01; ***p<0.001. Estimates derived from bivariate ANOVA models using survey weights and multiple imputation for missing data

**Table S8: Sex-specific Multivariable Poisson Regression Models for the association between high sodium consumption and sociodemographic, biomedical, and clinical characteristics.**

| Variables | Male (n=368) | | | Female (n=641) | | |
|---|---|---|---|---|---|---|
| | Prevalence Ratio | 95% CI | p-value | Prevalence Ratio | 95% CI | p-value |
| Age Category | | | | | | |
| *15-34 years* | Reference | | | | | |
| *35-54 years* | 1.11 | 0.92, 1.33 | 0.273 | 1.05 | 0.91, 1.21 | 0.480 |
| *55-74 years* | 0.93 | 0.75, 1.16 | 0.519 | 1.00 | 0.79, 1.27 | 0.971 |
| *≥ 75 years* | 0.74 | 0.57, 0.97 | 0.029 | 1.24 | 0.85, 1.81 | 0.273 |
| Blood Pressure Category | | | | | | |
| *Normal BP* | Reference | | | | | |
| *Prehypertension* | 0.94 | 0.80, 1.09 | 0.387 | 0.75 | 0.65, 0.86 | <0.001 |
| *Hypertension* | 1.07 | 0.91, 1.26 | 0.391 | 0.74 | 0.63, 0.87 | <0.001 |
| Blood Glucose Level | | | | | | |
| *Normal* | Reference | | | | | |
| *Impaired Fasting Glucose* | 1.03 | 0.88, 1.20 | 0.701 | 0.80 | 0.69, 0.94 | 0.006 |
| *(ADA) Diabetes* | 1.12 | 0.93, 1.35 | 0.240 | 0.96 | 0.80, 1.15 | 0.646 |
| BMI | | | | | | |
| *Normal (18.5 – 24.99 kg/m$^2$)* | Reference | | | | | |
| *Underweight (< 18.5 kg/m$^2$)* | 0.65 | 0.43, 0.97 | 0.033 | 0.89 | 0.47, 1.67 | 0.709 |
| *Overweight (25 – 29.99 kg/m$^2$)* | 1.05 | 0.90, 1.23 | 0.540 | 1.1093 | 0.89, 1.37 | 0.363 |
| *Obese (≥ 30 kg/m$^2$)* | 1.00 | 0.85, 1.18 | 0.998 | 1.47 | 1.27, 1.69 | <0.001 |
| Low GFR | 1.09 | 0.86, 1.37 | 0.480 | 0.62 | 0.44, 0.89 | 0.009 |
| Education Category | | | | | | |
| *Less than High School* | Reference | | | | | |
| *High School* | 1.07 | 0.93, 1.22 | 0.361 | 1.00 | 0.83, 1.20 | 0.974 |
| *More than High School* | 1.10 | 0.91, 1.33 | 0.330 | 0.97 | 0.78, 1.19 | 0.734 |
| House Possession Category | | | | | | |
| *0-8 items* | Reference | | | | | |
| *9-11 items* | 1.08 | 0.93, 1.22 | 0.246 | - | - | - |
| *12-22 items* | 1.02 | 0.85, 1.22 | 0.867 | - | - | - |
| Current Smoker (vs non-smoker) | 1.19 | 1.01, 1.40 | 0.038 | - | - | - |
| Physical Activity Level | | | | | | |
| *Low* | Reference | | | | | |
| *Moderate* | 1.08 | 0.89, 1.30 | 0.437 | 0.84 | 0.70, 1.01 | 0.058 |
| *High* | 0.93 | 0.78, 1.12 | 0.449 | 0.93 | 0.78, 1.10 | 0.382 |

Prevalence ratios derived from sex-specific multivariable Poisson regression models with high sodium consumption as outcome variable. Models were survey weighted and used multiple imputation. Smoking and household possession category were not included in models for females.

**Table S9: Sex-specific Multivariable Poisson Regression Models for the association between Low Potassium Consumption and sociodemographic, biomedical, and clinical characteristics.**

| Variables | Male (n=368) | | | Female (n=641) | | |
|---|---|---|---|---|---|---|
| | Prevalence Ratio | 95% CI | p-value | Prevalence Ratio | 95% CI | p-value |
| Age Category | | | | | | |
| *15-34 years* | Reference | | | | | |
| *35-54 years* | 1.54 | 1.00, 2.36 | 0.051 | 0.69 | 0.34, 1.38 | 0.290 |
| *55-74 years* | 0.64 | 0.35, 1.15 | 0.136 | 0.60 | 0.31, 1.17 | 0.132 |
| *≥ 75 years* | 0.49 | 0.26, 0.95 | 0.034 | 0.37 | 0.11, 1.25 | 0.108 |
| Blood Pressure Category | | | | | | |
| *Normal BP* | Reference | | | | | |
| *Prehypertension* | 0.51 | 0.30, 0.86 | 0.012 | 1.11 | 0.58, 2.13 | 0.754 |
| *Hypertension* | 1.27 | 0.72, 2.24 | 0.409 | 0.76 | 0.44, 1.30 | 0.306 |
| Blood Glucose Level | | | | | | |
| *Normal* | Reference | | | | | |
| *Impaired Fasting Glucose* | 1.10 | 0.70, 1.71 | 0.680 | 0.79 | 0.40, 1.54 | 0.484 |
| *(ADA) Diabetes* | 0.98 | 0.49, 1.97 | 0.963 | 1.22 | 0.61, 2.45 | 0.565 |
| BMI | | | | | | |
| *Normal (18.5 – 24.99 kg/m$^2$)* | Reference | | | | | |
| *Underweight (< 18.5 kg/m$^2$)* | 1.06 | 0.30, 3.72 | 0.928 | - | - | - |
| *Overweight (25 – 29.99 kg/m$^2$)* | 0.76 | 0.47, 1.23 | 0.256 | 2.81 | 1.38, 5.73 | 0.005 |
| *Obese (≥ 30 kg/m$^2$)* | 1.99 | 1.40, 2.82 | <0.001 | 4.50 | 2.32, 8.72 | 0.000 |
| High Cholesterol | 1.38 | 0.94, 2.04 | 0.100 | 2.09 | 1.12, 3.93 | 0.022 |
| Education Category | | | | | | |
| *Less than High School* | Reference | | | | | |
| *High School* | 0.46 | 0.30, 0.71 | 0.000 | 0.57 | 0.29, 1.12 | 0.102 |
| *More than High School* | 1.27 | 0.77, 2.01 | 0.343 | 0.88 | 0.44, 1.75 | 0.704 |

Prevalence ratios derived from sex-specific multivariable Poisson regression models with high sodium consumption as outcome variable. Models were survey weighted and used multiple imputation. Estimated prevalence ratio for females was 7.89 x e-09, hence these values were not included in the table.

**Table S10: Complete Cases Analysis: Sex-specific Multivariable ANOVA Models for the association between 24-hour urine sodium (log transformed) and sociodemographic, biomedical, and clinical characteristics.**

| | Male (n=208) | | | | Female (n=510) | | | |
|---|---|---|---|---|---|---|---|---|
| **Variables** | **β-coefficient** | **95% CI** | **p-value** | **% Change** | **β-coefficient** | **95% CI** | **p-value** | **% Change** |
| Age Category | | | | | | | | |
| *15-34 years* | Reference | | | | Reference | | | |
| *35-54 years* | -0.116 | -0.278, 0.046 | 0.161 | -10.9 | -0.096 | -0.241, 0.049 | 0.194 | -9.12 |
| *55-74 years* | -0.107 | -0.338, 0.125 | 0.363 | -10.1 | -0.267 | -0.523, -0.012 | 0.040 | -23.5 |
| *≥ 75 years* | -0.721 | -0.980, -0.462 | 0.000 | -51.4 | -0.240 | -0.657, 0.176 | 0.256 | -21.4 |
| Blood Pressure Category | | | | | | | | |
| *Normal BP* | Reference | | | | Reference | | | |
| *Prehypertension* | -0.445 | -0.597, -0.292 | 0.000 | -35.9 | -0.153 | -0.329, 0.024 | 0.089 | -14.1 |
| *Hypertension* | -0.107 | -0.286, 0.071 | 0.236 | -10.2 | -0.277 | -0.474, -0.080 | 0.006 | -24.2 |
| Blood Glucose Level | | | | | | | | |
| *Normal* | Reference | | | | Reference | | | |
| *Impaired Fasting Glucose* | -0.278 | -0.524, -0.032 | 0.027 | -24.3 | -0.137 | -0.252, -0.021 | 0.021 | -12.8 |
| *(ADA) Diabetes* | 0.165 | -0.108, 0.438 | 0.234 | 18.0 | 0.029 | -0.121, 0.178 | 0.705 | 2.90 |
| BMI | | | | | | | | |
| *Normal (18.5 – 24.99 kg/m²)* | Reference | | | | Reference | | | |
| *Underweight (< 18.5 kg/m²)* | -0.386 | -0.759, -0.012 | 0.043 | -32.0 | 0.332 | -0.348, 1.011 | 0.337 | 39.3 |
| *Overweight (25 – 29.99 kg/m²)* | 0.065 | -0.111, 0.240 | 0.466 | 6.68 | 0.124 | -0.069, 0.317 | 0.205 | 13.2 |
| *Obese (≥ 30 kg/m²)* | 0.564 | 0.348, 0.779 | 0.000 | 75.7 | 0.534 | 0.411, 0.658 | 0.000 | 70.6 |
| Low GFR | 0.145 | -0.233, 0.522 | 0.450 | 15.6 | -0.241 | -0.489, 0.007 | 0.057 | -21.4 |
| Education Category | | | | | | | | |
| Less than High School | Reference | | | | Reference | | | |
| High School | -0.114 | -0.294, 0.066 | 0.211 | -10.8 | -0.207 | -0.397, -0.017 | 0.033 | -18.7 |
| More than High School | 0.149 | -0.098, 0.395 | 0.236 | -16.0 | -0.376 | -0.616, -0.136 | 0.002 | -31.3 |
| House Possession Category | | | | | | | | |
| *0-8 items* | Reference | | | | Reference | | | |
| *9-11 items* | -0.164 | -0.322, -0.005 | 0.043 | -15.1 | - | - | - | - |
| *12-22 items* | -0.100 | -0.277, 0.078 | 0.268 | -9.49 | - | - | - | - |

| | Male (n=208) | | | | Female (n=510) | | | |
|---|---|---|---|---|---|---|---|---|
| **Variables** | **β-coefficient** | **95% CI** | **p-value** | **% Change** | **β-coefficient** | **95% CI** | **p-value** | **% Change** |
| Current Smoker (vs non-smoker) | 0.393 | 0.258, 0.529 | 0.000 | 48.2 | - | - | - | - |
| Physical Activity Level | | | | | | | | |
| *Low* | Reference | | | | Reference | | | |
| *Moderate* | 0.352 | 0.180, 0.525 | 0.000 | 42.2 | -0.105 | -0.284, 0.073 | 0.246 | -10.0 |
| *High* | 0.463 | 0.245, 0.680 | 0.000 | 58.8 | -0.052 | -0.162, 0.057 | 0.346 | -5.10 |

**Table S11: Complete Cases Analysis: Sex-specific Multivariable ANOVA Models for the association between 24-hour urine potassium (log transformed) and sociodemographic, biomedical, and clinical characteristics.**

| Variables | Male (n=208) | | | | Female (n=510) | | | |
|---|---|---|---|---|---|---|---|---|
| | β-coefficient | 95% CI | p-value | % Change | β-coefficient | 95% CI | p-value | % Change |
| Age Category | | | | | | | | |
| *15-34 years* | Reference | | | | | | | |
| *35-54 years* | 0.058 | -0.079, 0.194 | 0.405 | 5.93 | 0.114 | 0.002, 0.227 | 0.047 | 12.1 |
| *55-74 years* | 0.025 | -0.148, 0.199 | 0.773 | 2.57 | 0.026 | -0.067, 0.118 | 0.587 | 2.58 |
| *≥ 75 years* | -0.171 | -0.398, 0.057 | 0.139 | -15.7 | 0.046 | -0.116, 0.208 | 0.575 | 4.71 |
| Blood Pressure Category | | | | | | | | |
| *Normal BP* | Reference | | | | | | | |
| *Prehypertension* | -0.162 | -0.306, -0.018 | 0.027 | -15.0 | 0.083 | -0.054, 0.220 | 0.231 | 8.66 |
| *Hypertension* | -0.035 | -0.215, 0.144 | 0.697 | -3.48 | -0.010 | -0.126, 0.106 | 0.865 | -1.00 |
| Blood Glucose Level | | | | | | | | |
| *Normal* | Reference | | | | | | | |
| *Impaired Fasting Glucose* | 0.163 | 0.040, 0.286 | 0.010 | 17.7 | -0.027 | -0.126, 0.072 | 0.588 | -2.67 |
| *(ADA) Diabetes* | 0.265 | 0.110, 0.419 | 0.001 | 30.3 | 0.025 | -0.055, 0.105 | 0.543 | 2.49 |
| BMI | | | | | | | | |
| *Normal (18.5 – 24.99 kg/m²)* | Reference | | | | | | | |
| *Underweight (< 18.5 kg/m²)* | -0.128 | -0.379, 0.123 | 0.316 | -12.0 | 0.096 | -0.234, 0.426 | 0.565 | 10.1 |
| *Overweight (25 – 29.99 kg/m²)* | -0.074 | -0.187, 0.039 | 0.195 | -7.15 | 0.236 | 0.082, 0.391 | 0.003 | 26.6 |
| *Obese (≥ 30 kg/m²)* | 0.294 | 0.124, 0.464 | 0.001 | 34.2 | 0.462 | 0.328, 0.596 | 0.000 | 58.7 |
| High Cholesterol | 0.021 | -0.092, 0.134 | 0.710 | 2.15 | 0.239 | 0.159, 0.319 | 0.000 | 27.0 |
| Education Category | | | | | | | | |
| *Less than High School* | Reference | | | | | | | |
| *High School* | -0.156 | -0.252, -0.059 | 0.002 | -14.4 | -0.026 | -0.117, 0.066 | 0.584 | -2.51 |
| *More than High School* | -0.078 | -0.256, 0.010 | 0.388 | -7.49 | 0.119 | -0.021, 0.258 | 0.095 | 12.60 |
